# The CentiMarker Project: Standardizing Quantitative Alzheimer’s disease Fluid Biomarkers for Biologic Interpretation

**DOI:** 10.1101/2024.07.25.24311002

**Authors:** Guoqiao Wang, Yan Li, Chengjie Xiong, Yuchen Cao, Suzanne E. Schindler, Eric McDade, Kaj Blennow, Oskar Hansson, Jeffrey L. Dage, Clifford R. Jack, Charlotte E. Teunissen, Leslie M Shaw, Henrik Zetterberg, Laura Ibanez, Jigyasha Timsina, Cruchaga Carlos, the DIAN-TU Study Team, Randall J. Bateman

## Abstract

**Introduction:** Biomarkers have been essential to understanding Alzheimer’s disease (AD) pathogenesis, pathophysiology, progression, and treatment effects. However, each biomarker measure is a representation of the biological target, the assay used to measure it, and the variance of the assay. Thus, biomarker measures are difficult to compare without standardization, and the units and magnitude of effect relative to the disease are difficult to appreciate, even for experts. To facilitate quantitative comparisons of AD biomarkers in the context of biologic and treatment effects, we propose a biomarker standardization approach between normal ranges and maximum abnormal AD ranges, which we refer to as CentiMarker, similar to the Centiloid approach used in PET.

**Methods:** We developed a standardization scale that creates percentile values ranging from 0 for a normal population to 100 for the most abnormal measures across disease stages. We applied this scale to CSF and plasma biomarkers in autosomal dominant AD, assessing the distribution by estimated years from symptom onset, between biomarkers, and across cohorts. We then validated this approach in a large national sporadic AD cohort.

**Results:** We found the CentiMarker scale provided an easily interpretable metric of disease abnormality. The biologic changes, range, and distribution of several AD fluid biomarkers including amyloid-β, phospho-tau and other biomarkers, were comparable across disease stages in both early onset autosomal dominant and sporadic late onset AD.

**Discussion:** The CentiMarker scale offers a robust and versatile framework for the standardized biological comparison of AD biomarkers. Its broader adoption could facilitate biomarker reporting, allowing for more informed cross-study comparisons and contributing to accelerated therapeutic development.

## 1 Introduction

The quantification and comparison of Alzheimer’s disease (AD) biomarkers is crucial for understanding disease progression and the effectiveness of interventions. However, the current measures of fluid biomarker concentration, such as mass or moles per volume, are unique to each analyte, assay, and study.^1–4^ This lack of standardization makes it challenging to compare results across different studies, laboratories, and biomolecules. Therefore, standardization efforts are crucial to allow for quantitative comparison of the AD biomarkers in the context of both biologic and treatment effects.

One kind of standardization is an absolute reference standard for the molar amount of analyte per volume unit. Current practice and requirements by Health Authorities advise or require standardization of assays based on the metrology approach with traceability back to the SI-unit through certified reference materials (CRM) and reference measurement procedures for diagnostic biomarkers for clinical use, as has been done for cerebrospinal fluid (CSF) amyloid-beta 42 (Aβ42) by the International Federation of Clinical Chemistry (IFCC) Work Group.^5^ The final Aβ42 CRMs have also been used for re-calibrating commercial immunoassays for CSF Aβ42, including those that are FDA approved.^6^ However, this approach is very laborious and time-consuming.

Other standardization methods, such as z-score normalization, have been employed to facilitate comparisons of biomarkers within or across studies.^7^ Z-score normalization typically involves utilizing a "control" group, such as a baseline or a reference group, like young healthy controls, to convert raw values or log-transformed values into z-scores. In this case, 1 unit change in z-score represents 1 SD change in the original scale. In our study, we aim to introduce an alternative, the concept of CentiMarkers, which is designed to transform fluid biomarker values onto a scale from 0 to 100. CentiMarkers aim to provide a common metric that allows for quantitative comparisons of AD biomarkers between normal and near maximum abnormal ranges. This would be helpful for comparing the same analyte and assay across different studies and cohorts. Additionally, this would be helpful in comparing fluid biomarker values across different assays measuring the same analytes. For instance, the measurement of amyloid-β 42/40 using different antibodies, immunoassays, or mass spectrometry methods can be standardized, enabling meaningful comparisons both within and across studies.

Recent evidence from interventional trials suggests that some AD biomarkers that reach normal levels, such as achieving amyloid PET negativity, have predictive value for clinical benefit.^2,3,8^ Additionally, other biomarkers such as p-tau217 may be associated with cognitive impairment and response to treatment.^9^ However, comparing different disease cohorts, various forms of the disease, and diverse populations is often impractical due to the lack of a standardized biologic metric for fluid biomarker measurements. Having a common metric for these diverse analytes enables the interpretation of their biologic effects in relation to disease progression or response to interventions. Tracking disease progression or modification often involves monitoring multiple biomarkers, such as amyloid, tau, and neurodegeneration markers. The different AD biomarkers change sequentially over a 30-year disease span (20 years before symptom onset through 10 years after symptom onset) and may not be monotonic.^10–13^ Having a common metric for these diverse analytes enables the interpretation of their biologic effects in relation to disease progression or response to interventions.

The objectives of this study are the following: (i) to propose a mechanism for calculating CentiMarkers and showcase its applicability in different study cohorts; and (ii) to illustrate the usefulness of CentiMarkers values in facilitating the comparison of treatment effects across various fluid biomarkers for dominantly inherited AD (DIAD). This publication details the scope of use, methodology, contrasts approach for other purposes, addresses potential limitations, and explores alternative approaches. Future work will focus on how to generalize and apply the CentiMarker approach for other studies and purposes.

## 2 Methods

### 2.1 Study Oversight

The DIAN-TU study was conducted following the principles outlined in the Declaration of Helsinki and adhered to the guidelines set by the International Council for Harmonization and Good Clinical Practice. Ethical approval from the respective ethics committees at each participating site was obtained. Prior to participating in the study, all individuals provided written informed consent.

### 2.2 Study Participants

DIAN-TU-001 is a randomized, placebo-controlled, multi-arm trial of gantenerumab or solanezumab in participants with DIAD across asymptomatic and symptomatic disease stages (NCT01760005). Mutation carriers were assigned 3:1 to either drug or placebo and received treatment for 4–7 years. The information about these participants as well as the dosing schedule has been published in previous studies.^1,4,8^ Briefly, the trial included 193 participants, consisting of 144 mutation carriers (MCs) and 49 non-mutation carriers (NCs). The participants in the trial were either cognitively normal (Clinical Dementia Rating® [CDR = 0]) or had early-stage disease (CDR 0.5 or 1, indicating very mild or mild dementia) at the time of enrollment. The DIAN-TU trial presents a unique opportunity to utilize a subject set of young to middle-aged healthy controls. These controls consist of family members of mutation carriers without mutations (referred to as non-carriers or NCs). These NCs are young, healthy individuals without AD-related disease pathologies.

ADNI is a longitudinal study conducted in the United States focusing on sporadic Alzheimer’s disease and individuals at risk who undergo neuroimaging and biomarker evaluations. The study data utilized for our analysis ranged from 2008 to 2022. We specifically selected a subset of ADNI participants with derived estimated years from symptom onset (EYO), as reported in a previous study.^14^ Two fluid biomarkers, which are common to both the DIAN-TU-001 and ADNI studies, were converted into CentiMarkers to facilitate illustration and comparison.

### 2.3 Statistical Methods

The parameters for calculating CentiMarker values in each of the two cohorts (DIAN-TU and ADNI) were established using their respective data. Subsequently, in the re-analysis of the DIAN-TU-001 trial, we interpreted the treatment effect of gantenerumab based on the CentiMarker unit, which provided a meaningful measure for assessing the efficacy across different fluid biomarkers.

#### 2.3.1 CentiMarker Calculation

In order to convert the raw biomarker values to CentiMarker values, we utilized a similar methodology as the Centiloid conversion approach.^15^ For CentiMarker calculations, more abnormal biomarker raw values indicate worse disease stages as defined by the direction of disease vs. normal groups. In summary, the process involves the following steps:

1. Identification of the CentiMarker-0 data set and determination of the CM-0 CentiMarker anchor value based on the CentiMarker-0 cohort.
2. Identification of the CentiMarker-100 data set and determination of the CM-100 CentiMarker anchor value based on the CentiMarker-100 cohort.
3. Calculation of the CentiMarkers.

#### Identification of the CentiMarker-0 data set

To establish the CentiMarker-0 anchor (µ_CM-0_), all the data from the asymptomatic mutation non-carriers enrolled in the DIAN-TU-001 trial is utilized. The first step involves calculating the interquartile range (IQR) of the data. Then, any outliers that fall outside the range of (> Q3 + 1.5 X !QR) or (< Q1- 1.5 X !QR) are excluded from the dataset.^15^ Finally, the mean of the remaining data points, was established as the CentiMarker-0 anchor for subsequent CentiMarker calculations.

#### Identification of the CentiMarker-100 data set

To determine the CentiMarker-100 anchor (γ_CM-100_), data from all mutation carriers enrolled in the DIAN-TU trial who did not receive active treatments were used. This includes both the baseline data from participants who were assigned to the active treatment as well as the baseline and post-baseline data from those who were assigned to placebo.

The same approach applied in the identification of the CentiMarker-0 data set, is again employed to identify and remove outliers from this dataset. The CentiMarker-100 anchor, denoted as y_MC-100_, is established as the 95th percentile of the most abnormal value across the spectrum of disease stages in the CentiMarker-100 dataset. With this approach, when the higher values indicate more severe disease stage, the 95^th^ percentile of the most abnormal value corresponds to the 95^th^ percentile of the highest values; when the lower values indicate more severe disease stage, the 95^th^ percentile of the most abnormal value corresponds to the 5^th^ percentile of the lowest values. The bootstrapping method was used to calculate the standard deviation of the 95^th^ percentile of the most abnormal value. When a biomarker demonstrates a monotonic disease progression trajectory, higher CentiMarkers consistently indicate a more severe disease stage, irrespective of the direction of its raw values. However, for biomarkers that do not exhibit a monotonic decline (e.g., decreasing after reaching a certain disease stage), higher values of the CentiMarker do not necessarily correspond to a more severe disease stage.

#### Calculation of CentiMarkers

The CentiMarker is calculated as:

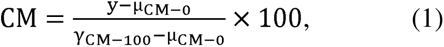

where y is the raw biomarker value, µ_CM-0_ is the CentiMarker-0 anchor, γ_CM-100_ is the CentiMarker-100 anchor

#### 2.3.2 Benefit of Using the Same CentiMarker-0 Anchor Cohort

Due to the two-anchor approach involved in CentiMarker conversion, namely the 0 and 100 value, maintaining consistency in the CentiMarker-0 subject set is crucial. This not only facilitates the comparability of CentiMarker calculated using different CentiMarker-100 anchors, but also makes them exchangeable. By keeping the CentiMarker-0 cohort constant, we can utilize the same CentiMarker-0 anchor (µ_CM-0_) when calculating CentiMarkers values regardless how the γ_CM-100_ is determined. Assuming the CentiMarkers is calculated by two different CentiMarker-100 anchors as following:

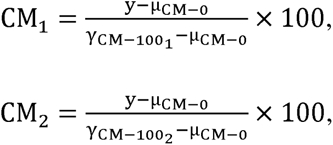

where γ_CM-1001_ and γ_CM-1002_ are the two different CentiMarker-100 anchors. Rearranging these two formulas, we have:

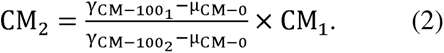

Therefore, by using the same CentiMarker-0 cohort, the method to determine the γ_CM-100_ is inherently exchangeable. In this study, we selected the 95^th^ percentile of the most abnormal value as the CentiMarker-100. CentiMarkers obtained through alternative methods, such as using the 90^th^ percentile of the most abnormal value or the mean of all mutation carriers, can be converted into each other using formula (2).

## 3 Results

### 3.1 Using the 95 Percentile of the Most Abnormal Value as the CentiMarker-100 Anchor

The two anchor values, CentiMarker 0 and CentiMarker 100, which were established using the DIAN-TU-001 data for each biomarker, are provided in Table 2. These values were then utilized in formula (1) to calculate the CentiMarkers for each biomarker accordingly.

**Table 1:** Baseline demographics for each cohort.

|  | DIAN-TU 001 mutation carriers |  |  | ADNI |  |  |
| --- | --- | --- | --- | --- | --- | --- |
|  | Active<br>gantenerumab<br>N=52 | Active<br>solanezumab<br>N=50 | Active<br>placebo<br>N=40 | Controls<br>N=69 | MCI group<br>N=335 | AD group<br>N=160 |
| Age (years) | 46.0 ± 10.8 | 42.5 ± 9.5 | 44.2 ± 9.6 | 74.9 ± 5.4 | 73.3 ± 7.2 | 73.9 ± 8.0 |
| Female (n(%)) | 21 (40) | 29 (58) | 22 (55) | 37 (54) | 130 (39) | 67 (42) |
| Education (years) | 14.8 ± 3.1 | 14.9 ± 2.9 | 15.5 ± 3.1 | 16.2 ± 2.6 | 15.9 ± 2.8 | 15.7 ± 2.7 |
| CDR 0 (n (%)) | 31 (60) | 30 (60) | 22 (55) | 69 (100) | 1 (0.3) | NA |
| CDR 0.5 (n (%)) | 15 (29) | 13 (26) | 15 (38) | NA | 334 (100) | 68 (43) |
| CDR 1 (n (%)) | 6 (12) | 7 (14) | 3 (8) | NA | NA | 91 (57) |
| CDR >= 2 (n (%)) | NA | NA | NA | NA | NA | 1 (0.6) |
| CDR-SB (n (%)) | 1.33 ± 2.08 | 1.37 ± 2.01 | 1.43 ± 1.87 | 0.07 ± 0.17 | 1.62 ± 0.89 | 4.52 ± 1.64 |
| MMSE | 27.10 ± 3.45 | 26.72 ± 4.11 | 26.68 ± 3.97 | 29.09 ± 1.04 | 27.58 ± 1.76 | 23.14 ± 2.09 |

**Table 2:** CentiMarker-0 and CentiMarker-100 values for each biomarker within the DIAN-TU- 001 population.

| Biomarker | CentiMarker 0 values<br>(Mean ± SD (n)) of NMCs | CentiMarker 100 values (95 Percentile of<br>the Most Abnormal Score ± SD (n)) of MCs |
| --- | --- | --- |
| NTK CSF Tau (pg/mL) | 135.20 ± 30.87 (109) | 399.90 ± 7.68 (197) |
| NTK CSF p-Tau181 (pg/mL) | 11.95 ± 2.45 (93) | 52.93 ± 1.41 (194) |
| NTK CSF Neurofilament Light<br>Chain Protein (pg/mL) | 67.69 ± 20.33 (113) | 242.60 ± 10.27 (203) |
| NTK CSF Neurogranin<br>(pg/mL) | 842.82 ± 223.63 (119) | 2066.00 ± 68.77 (205) |
| NTK CSF YKL-40 Protein<br>(ng/mL) | 105.35 ± 33.71 (117) | 247.20 ± 8.71 (205) |
| NTK CSF Glial Fibrillary<br>Acidic Protein (ng/mL) | 5.14 ± 1.88 (116) | 13.75 ± 0.80 (209) |

Figure 2 illustrates the CentiMarkers for some of the fluid biomarkers published within the DIAN-TU-001 trial.^16^ As anticipated, most CentiMarkers are observed to fall within the range of 0 to 100 and demonstrate an increasing trend as the disease progresses. Table 3 presents the means (SD) of CentiMarkers for both the CentiMarker-0 and CentiMarker-100 datasets. As anticipated, the means for the CM-0 dataset are all zero. In contrast, the means for the CentiMarker-100 dataset change as the disease advances from asymptomatic (CDR 0) to symptomatic (CDR>0). Note that not all measures go up, and some measures are not monotonic, as they increase and then decrease as the disease advances (e.g., CSF tau).^11,17^ By utilizing CentiMarkers in this way, the maximum and minimum abnormalities of the biomarker measures can be expressed on a common and simple scale to enable easy interpretation.^11,18^

**Figure 1:**
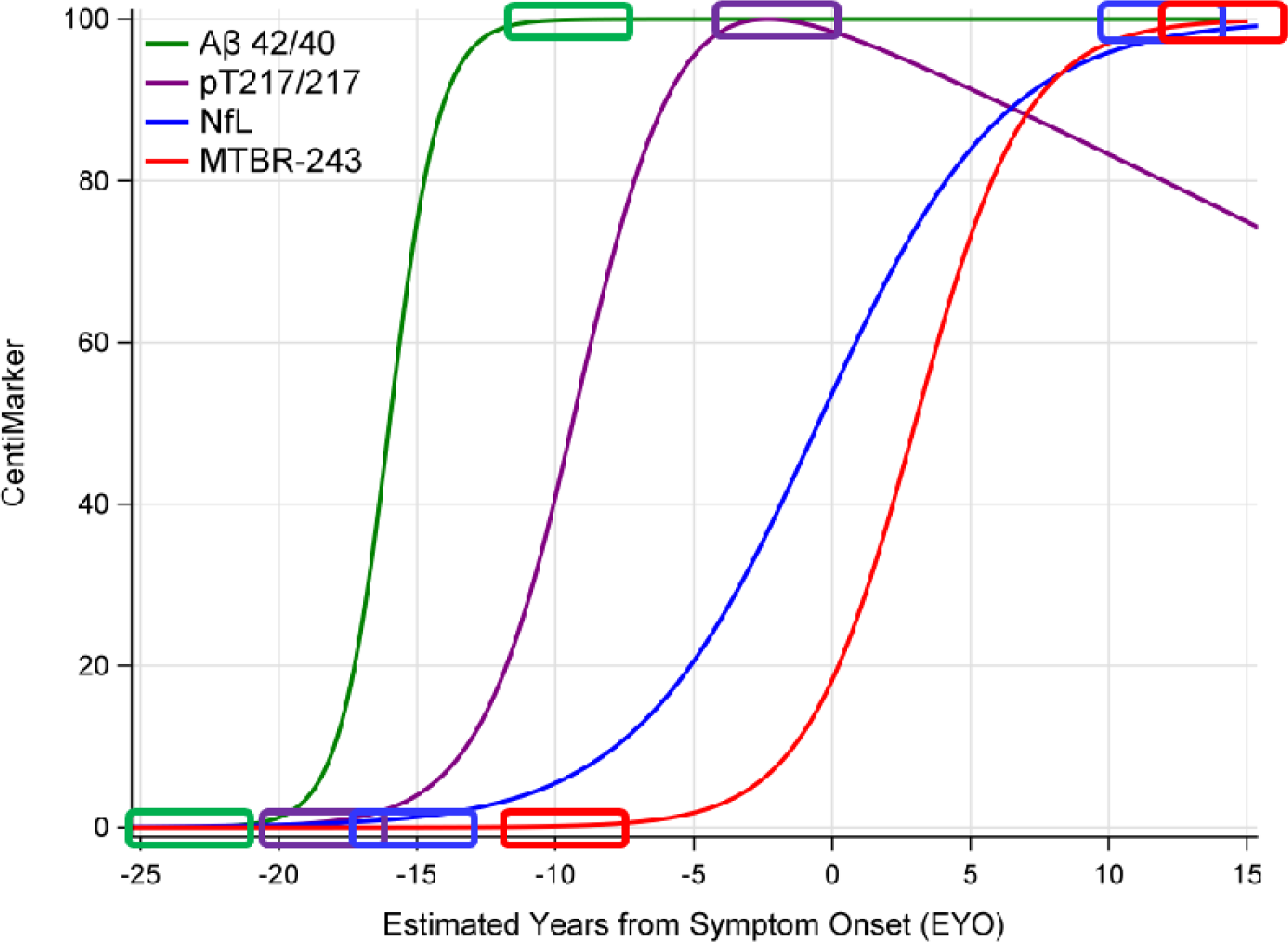
Illustration of CentiMarkers Approach. The rectangles indicate the CentiMarker 0 and CentiMarker 100 for each biomarker Figure 1 illustrates the disease progression trajectories utilizing the CentiMarker concept. In this approach, each fluid biomarker is rescaled to a range of 0 to 100, enabling a straightforward, intuitive, and direct comparison across different biomarkers. Additionally, Table 1 provides the baseline demographics of all participants included in this study for each respective cohort.

**Figure 2:**
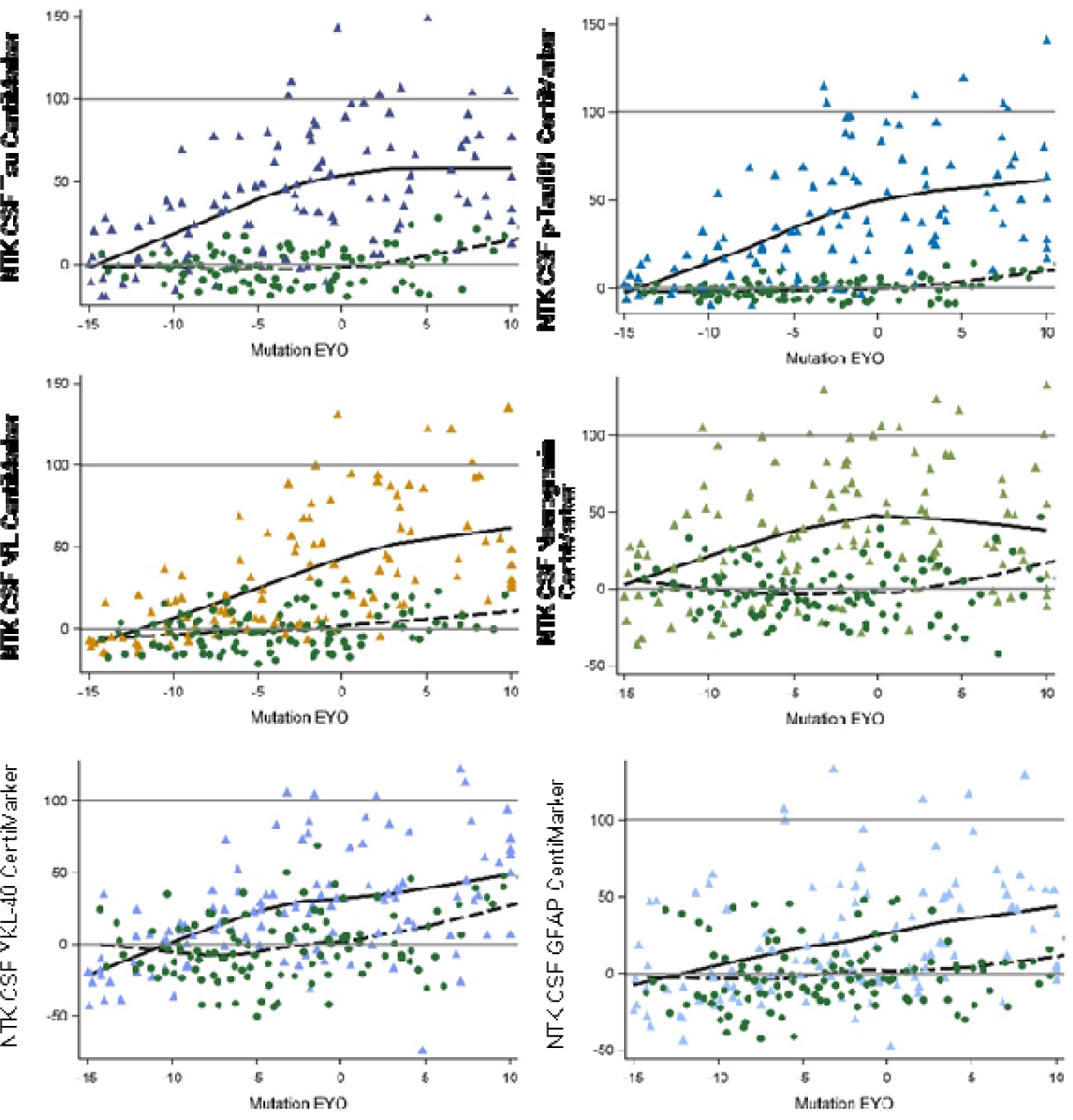
Comparison of different CentiMarkers in the same study by stage of disease comparing normal controls (green circles) to mutation carriers (color triangles). The mutation carriers included only the baseline data from the treatment groups and all the data from the placebo group in the DIAN-TU-001 trial. The x-axis represents the Estimated Years to Symptom Onset (EYO) based on mutation information, with zero indicating the expected onset of symptoms, covering a range of 25 years to represent disease progression.

**Table 3:** Mean ± SD (N) of CentiMarkers for the CentiMarker-0 dataset and CentiMarker-100 dataset by CDR global for the DIAN-TU-001 cohort.

| **Biomarker** | CM-0 (NMCs),<br>mean±SD (N) | CM-100 (MCs), mean±SD (N) | | | |
| --- | --- | --- | --- | --- | --- |
|  | **CDR = 0** | **CDR = 0** | **CDR = 0.5** | **CDR = 1** | **CDR ≥ 2** |
| NTK CSF Tau (pg/mL) | 0.00 ± 11.66 (109) | 25.31 ± 28.50 (119) | 56.72 ± 34.53 (55) | 68.39 ± 32.60 (18) | 42.37 ± 49.43 (5) |
| NTK CSF p-Tau181 (pg/mL) | 0.00 ± 5.98 (93) | 22.95 ± 26.73 (116) | 52.84 ± 30.48 (54) | 68.82 ± 33.99 (19) | 28.50 ± 35.27 (5) |
| NTK CSF neurofilament light chain (Log) | 0.00 ± 23.18 (118) | 14.55 ± 27.81 (123) | 56.64 ± 24.32 (63) | 82.65 ± 28.26 (21) | 76.60 ± 31.46 (5) |
| NTK CSF Neurogranin (pg/mL) | 0.00 ± 18.28 (119) | 24.77 ± 35.08 (123) | 45.23 ± 36.61 (59) | 52.11 ± 32.45 (18) | 11.74 ± 50.52 (5) |
| NTK CSF YKL-40 Protein (ng/mL) | 0.00 ± 23.77 (117) | 13.04 ± 31.27 (122) | 36.87 ± 37.88 (60) | 60.50 ± 38.72 (18) | 50.86 ± 45.53 (5) |
| NTK CSF Glial Fibrillary Acidic Protein (ng/mL) | 0.00 ± 21.81 (116) | 9.50 ± 32.89 (123) | 35.04 ± 39.92 (61) | 37.15 ± 28.95 (20) | 38.23 ± 58.91 (5) |

### 3.2 Treatment Effect in CentiMarkers

Figure 3 illustrates the change from baseline to year 4 for both the gantenerumab and the placebo groups as measured in both CentiMarker units and in raw values (as previously published^16^).

**Figure 3:**
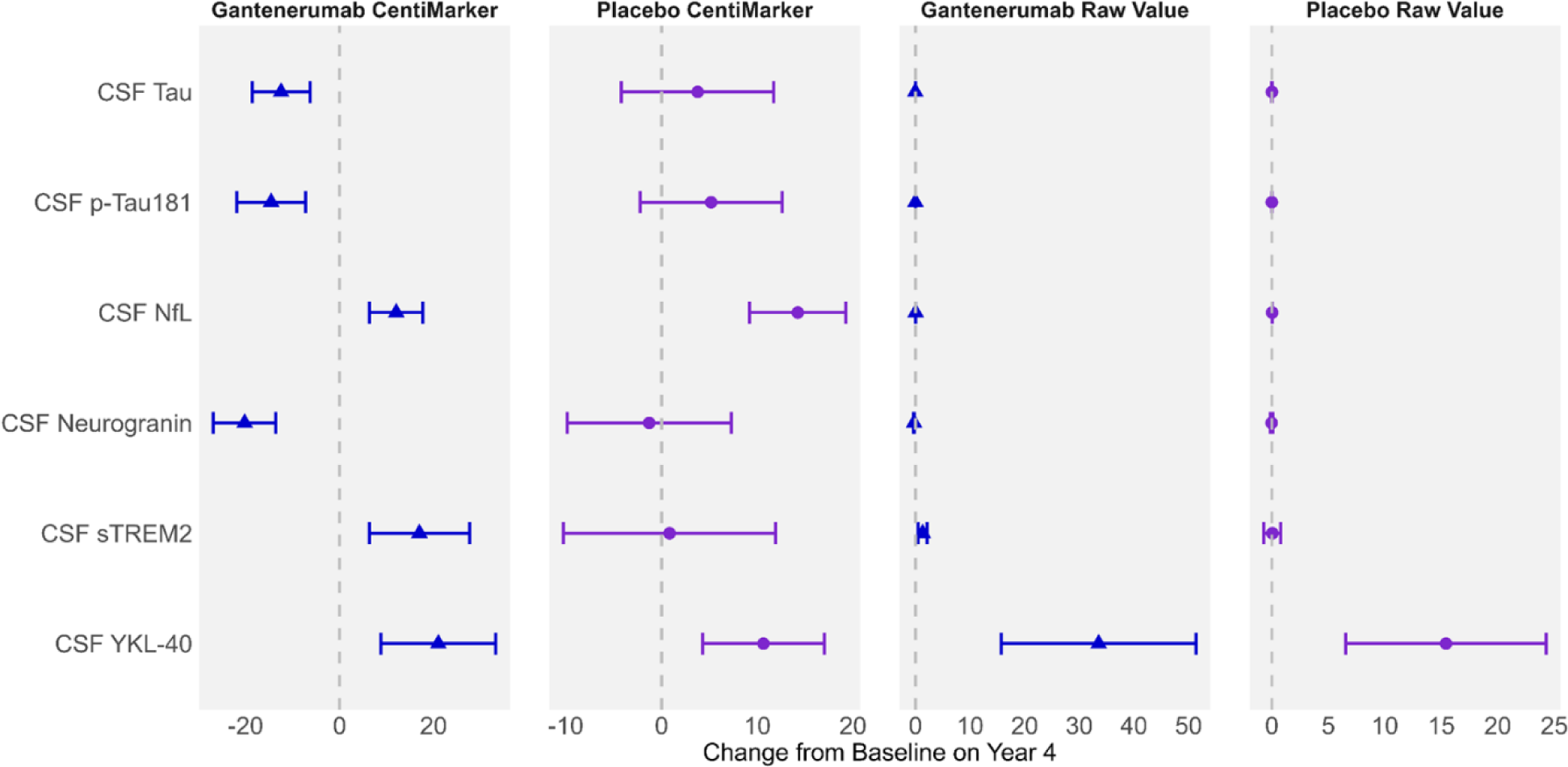
Utilizing CentiMarkers facilitates the interpretation and comparison across biomarkers by converting them to a similar scale, while using raw values is more difficult to compare as units are different and not scaled to disease ranges. Estimated mean change from baseline in CentiMarkers with 95% confidence intervals for the gantenerumab and placebo groups using MMRM analyses in the DIAN-TU-001 trial. These results demonstrate the magnitude of disease normalization compared to normal (CM 0) vs. fully abnormal (CM 100) states. Raw values are in the unit of ng/mL

CentiMarkers facilitate the interpretation and comparison of the treatment effect compared to the raw values. The change from baseline to year 4 in both the gantenerumab and the placebo groups are comparable in CentiMarkers across all the biomarkers, which contrasts with those using raw values.^16^ Supplemental Figure 1 illustrates the comparison, including GFAP. It is worth noting that GFAP exhibits larger variability in the gantenerumab group, making it less comparable to other biomarkers. However, the change observed in the placebo group remains consistent across all biomarkers, including GFAP.

Supplemental Figure 2 illustrates the change from baseline over a 4-year treatment period. The CentiMarkers clearly depict the distance of participants from the normal level at baseline and the extent to which the treatment has brought them closer to the normal level after 4 years. For instance, when considering CSF tau, the administration of gantenerumab resulted in a decrease of 12 units in CentiMarker, reducing it from 46 to 34. However, it is important to note that even at the end of the treatment, participants still remained 34 units above the normal CentiMarker value.

Figure 4 illustrates the comparison of disease progression for each biomarker across a time span of 15 years preceding symptom onset to 10 years following symptom onset. This comparison included three groups: mutation non-carriers (i.e. cognitively normal individuals), mutation carriers treated with gantenerumab, and mutation carriers not receiving gantenerumab treatment. CentiMarkers allow us to quickly identify biomarkers that exhibit a similar pattern of progression and evolve at a similar rate.

**Figure 4:**
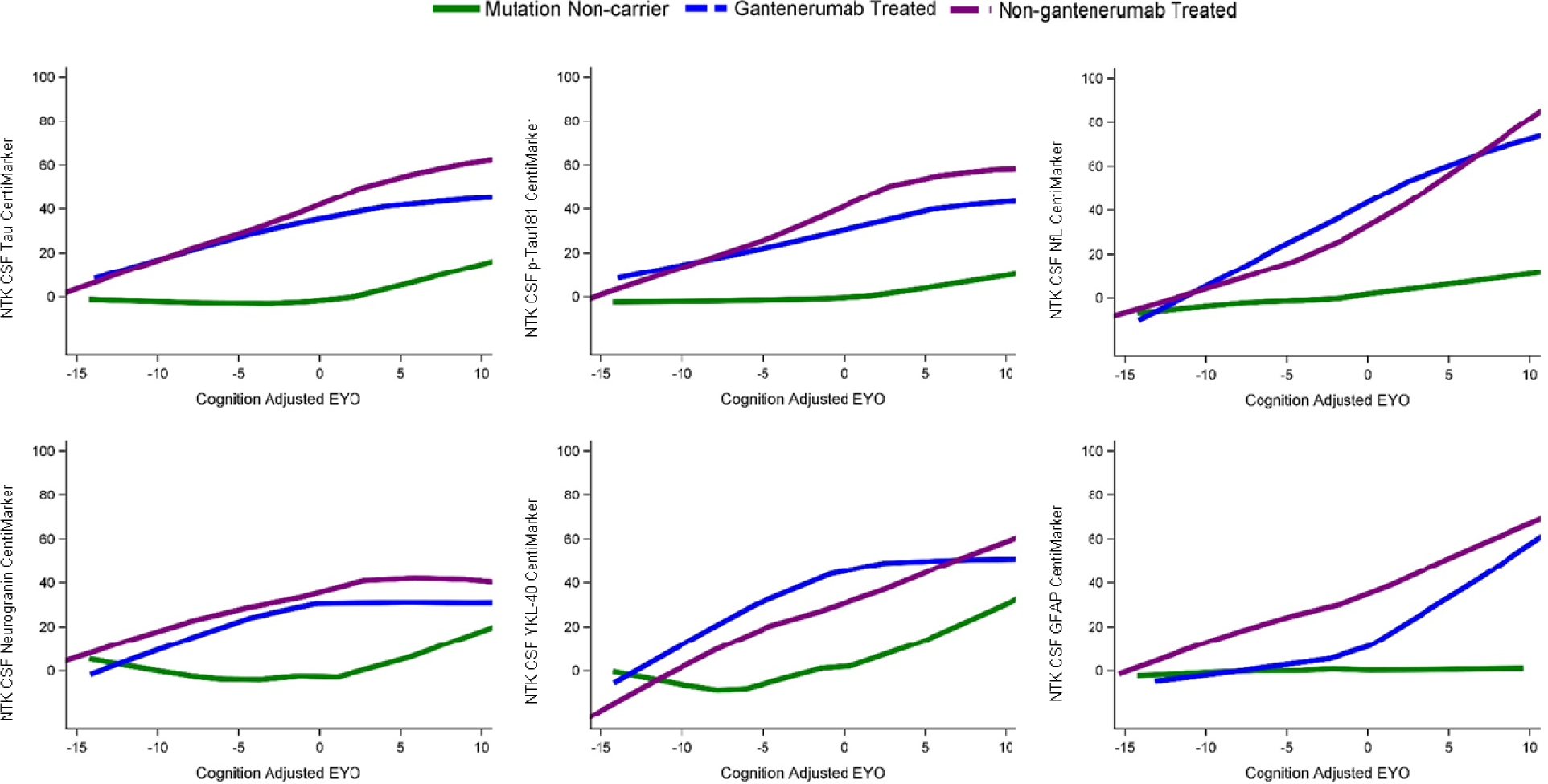
Illustration of treatment effects over EYO relative to mutation non-carriers (green line). Participants treated with gantenerumab (blue line) showed slower progression than those non-gantenerumab treated (purple line) across multiple biomarkers. The relative differences of treatment effect indicate that although there were improvements in multiple biomarkers, the measures did not reach normal CentiMarker levels for some measures.

**Figure 5:**
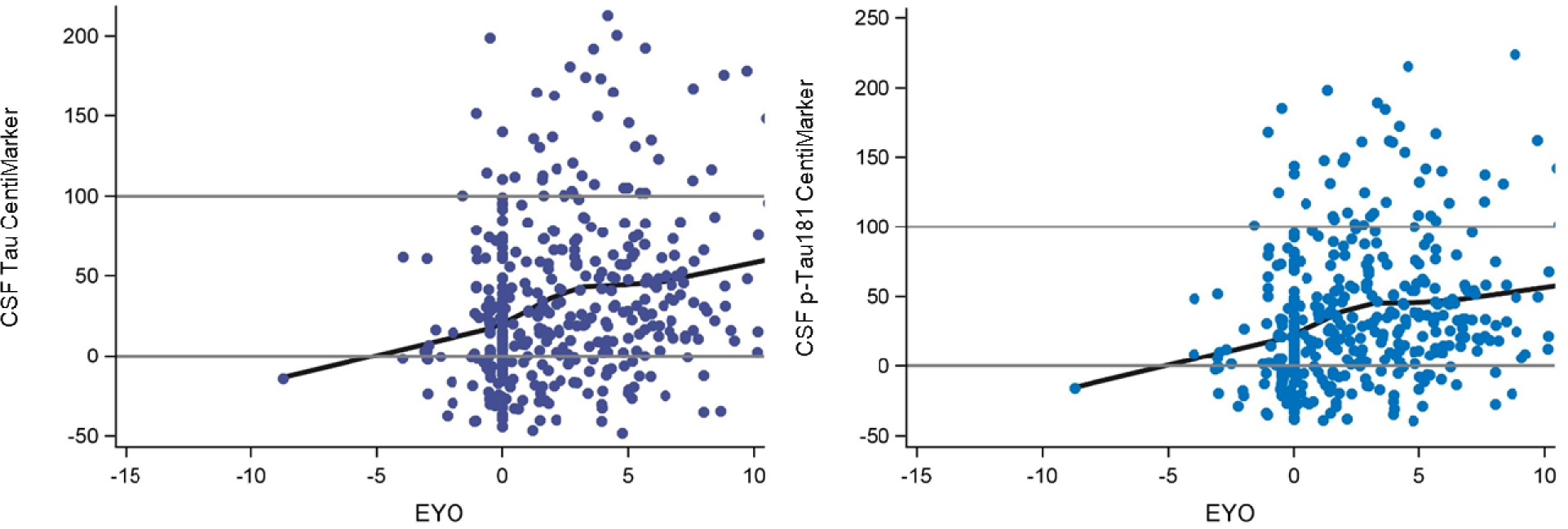
Illustration of CentiMarkers in ADNI. EYO is calculated by using the individual’s own age of symptom onset as EYO of 0. These distributions replicate increasing CSF tau and p- tau181 by stage of disease.

### 3.3 CentiMarkers in ADNI

In the ADNI study, the calculation of CentiMarker 0 (normal cohort) incorporated datasets from the clinical normal cohort. This cohort was defined as individuals with a CDR score of 0, indicating no significant cognitive impairment. Additionally, participants in this cohort had a diagnosis status of cognitive normal and fell within the age range of 50 to 89 years old. The CentiMarker 100 cohort in the study consisted of individuals with cognitive impairment, specifically those with a CDR score greater than 0. This cohort included individuals diagnosed with mild/moderate cognitive impairment or AD and had an age range of 50 to 89 years old. The CentiMarker 0 and CentiMarker 100 for CSF total tau are 240.03 ± (80.76) and 527.50 (8.44); and 21.57 ± (7.65) and 54.28 (1.03) for CSF p-tau181. Figure 6 illustrates the CentiMarkers of the two shared fluid biomarkers collected in both the DIAN-TU and ADNI studies. Similar to the findings in the DIAN-TU studies, the CentiMarkers in the ADNI study mostly fall below 100 and tend to cluster within the range of 0 to 100.

## 4 Discussion

AD researchers have developed biomarkers of AD pathogenesis, pathophysiology, progression, and treatment effects that have become amongst the most advanced in both breadth and accuracy of common diseases. However, because each biomarker measure is a representation of the biological target, the assay used to measure it, and the variance of the assay, these biomarker measures are difficult to compare between biomarkers, across studies, and even within a biomarker type (e.g., amyloid-β or phospho-tau concentrations). To facilitate quantitative biological comparison of AD biomarkers in the context of biologic and treatment effects, in this paper we propose an approach, which we refer to as CentiMarker, to re-scale biomarker levels for standardization between normal ranges and maximum abnormal AD ranges similar to the Centiloid approach used for amyloid PET. Other standardization efforts are focused on reference standards to make raw values of the same biological measure comparable across labs (e.g., IFCC standardization), or harmonizing one measure across assay run and labs per Good Laboratory Practices (GLP). These standardization projects typically take years to complete, and some of the biomarkers may be hard to standardize due to molecular heterogeneity of the analyte to be standardized, which may result in non-commutable reference materials.^19^ In contrast, the goal of the CentiMarker standardization is to have a biologically relevant scale that relates to the disease stage from normal to abnormal to facilitate biologic interpretation. Thus, these multiple standardization approaches are useful in different contexts: comparing the same measure across labs, harmonizing across runs and labs, and comparing the magnitude of biological effects across biomarker measures in an easy-to-understand scale.

AD is a complex neurodegenerative disease characterized by the accumulation of amyloid-beta plaques and tau tangles in the brain with associated inflammation and neuronal injury. These pathological changes are often accompanied by measurable brain atrophy and cognitive decline. However, the relationship between the magnitude of change in biomarkers and the clinical symptom of AD is not linear, making it challenging to interpret the disease progression and the effects of potential interventions.

### Rationale for establishing CentiMarkers

We have developed a conceptual framework to facilitate the interpretation of biomarker changes in AD studies. This approach, called CentiMarkers, aims to mathematically transform biomarker measurements onto a standardized scale, with 0 representing normal and 100 representing nearly (95%) maximally abnormal levels, conceptionally similar but not identical to Centiloid scales used in PET imaging.^15^ This transformation addresses several challenges in AD biomarker research.

Firstly, it provides a more intuitive measure of the magnitude of biomarker change. This is essential for both observational studies and clinical trials, facilitating comparisons between drugs and their effects at different stages of the disease and the comparison of stage of disease across cohorts. Secondly, this approach partly addresses the lack of comparability across biomarker assays. While no standardization can provide cross-assay equivalence, it enables a reasonable approximation for understanding overall trends in biomarker dynamics. Importantly, this concept was developed through extensive discussion and collaboration with experts across research groups in biomarkers, imaging, clinical studies, and interventional trials. This manuscript introduces the CentiMarker method, offering a conceptual framework and practical application for its use in observational studies and interventional trials. The method is demonstrated using large AD cohorts from the ADNI and DIAN-TU-001 studies, providing real-world examples. We anticipate that this approach will enhance the communication of AD biomarker results, making them more readily understood by the broader research community. Wide adoption of this approach could lead to improved comparability between studies and potentially aiding in a more unified understanding of AD pathophysiology as reflected by biomarkers.

### Defining normal and abnormal ranges for normalization

The CentiMarker 0 is the mean value in a normal cohort, defined by each study. The CentiMarker approach uses the 95^th^ percentile most abnormal value (i.e., the value defining the most abnormal 5% of the affected population) to define the CentiMarker 100, which is different from amyloid PET Centiloid, which uses the mean of individuals with early symptomatic AD to define Centiloid 100 (50% percentile). Unlike amyloid PET, which has a monotonic change until very late in the course of AD, fluid biomarkers may reach their maximum earlier in the course of AD, and importantly reach maximums at different stages of disease (see Figure 1). Therefore, CentiMarker 100 is defined by the 95^th^ percentile because of the more complex trajectories of these biomarkers. By establishing CentiMarker 100 as the upper abnormal value for biomarkers across the entire range of disease stages and time, a maximum abnormal upper range is set, irrespective of when and how the biomarker changes or whether it exhibits non-monotonic behavior. In cases where only a small cohort is available for deriving the CentiMarker 100 value or concerns exist regarding non-representative outliers, an alternative approach can be implemented. Rather than relying on the maximum upper range, the CentiMarker 100 value can be determined using either the 90^th^ percentile abnormal value within the available cohort or by referencing the CentiMarker 100 value reported in other studies utilizing the same assay.

To derived the CentiMarker scale,^15^ we recommend having a minimum of 30 data points for the CM 0 group in order to obtain a relatively accurate estimation of the mean. Through extensive bootstrapping estimation, our analysis suggests that having 30 to 50 data points for the CM 100 groups can yield a relatively stable value for the 95th most abnormal value. Notably, 50% to 70% of the bootstrapped 95th most abnormal values fall within 10% of the overall group’s 95% most abnormal value, varying depending on the biomarkers’ variability.

Another issue is that as the disease advances, some research participants become too impaired to continue in research studies. Some biomarkers (e.g., amyloid PET) may plateau, but other biomarkers (e.g., brain atrophy) likely continue to progress even after participants are too impaired to participate in studies, thus precluding defining the full disease course for CM 100.

Future work would need to address this for studies that extend beyond the defining CM 0 and 100 groups.

### Use of CentiMarkers

CentiMarkers offer a solution for ensuring comparability across different biomarker measures and units, as well as accounting for disease effects. By transforming biomarker measurements onto a common scale ranging from normal to maximally abnormal, CentiMarkers provide a standardized framework for accurate comparison and assessment. This allows for a more intuitive understanding of the magnitude of biomarker changes and their relationship with the clinical symptoms of AD. For instance, a CentiMarker value of 50 would indicate that a biomarker is halfway between the normal and maximum abnormal ranges, providing a clear and straightforward interpretation of the biomarker status. Moreover, CentiMarkers can be used to compare the effects of different interventions on AD biomarkers.

This is particularly important in the context of clinical trials, where multiple biomarkers are often assessed simultaneously. For instance, let’s consider a scenario where a CentiMarker originally measured 50 and after treatment, it decreased to 0. In this case, the biological measure abnormality has been effectively corrected. On the other hand, if the CentiMarker decreased from 100 to 50, despite a similar magnitude of change, the measure has not been biologically corrected. By providing a common metric for all biomarkers, CentiMarkers allow us to differentiate between treatments that fully normalize the biological measure and those that only produce partial improvement, and also facilitate direct comparisons of the effects of different biological effects.

### Future Research and Application

The application of CentiMarkers is not limited to AD research. The concept can be generalized to other neurodegenerative diseases, such as Parkinson’s disease and Huntington’s disease, and even to other fields of medicine, such as cardiovascular disease, cancer, and infectious diseases. This broad applicability makes CentiMarkers a potentially powerful tool for effective communication of biomedical research results.

### Challenges and Limitations

However, the implementation of CentiMarkers also has its challenges. It requires a well-characterized disease cohort and a normal control to define CentiMarkers. Each assay must be tested on these cohorts, which can be resource intensive. Furthermore, different ways to choose maximum and minimum abnormality may lead to different scales, limiting comparisons. Other harmonization techniques, like the use of z-scores, have been employed to facilitate direct comparisons across fluid biomarkers. However, it is important to note that the z-score approach does not incorporate disease normality into its scale. Additionally, a one-unit change in z-score corresponds to a one standard deviation (SD) change in the original raw values. Therefore, the comparability of z-scores relies on measurement variability, which may be influenced by factors such as assay precision and biological variation. Other efforts utilize certified reference standards to make individual assays or analytes comparable across studies, but this only works within an analyte, not across analytes, and again does not provide relative disease information.

Future research directions include testing the approach in more studies, cohorts, and individual biomarkers to validate and refine the method.^7^ Additionally, collaboration among various research groups will be crucial in establishing bridging cohorts similar to the Centiloid’s approach.^20^ This collaborative effort aims to facilitate the conversion of CentiMarker measurements across various cohorts and assays, thereby enhancing comparability across different fluid biomarkers. An important future goal and challenge will be to ensure researcher understanding, uptake, and utilization of CentiMarkers in their studies. The coordination and harmonization of specific approaches are necessary to enable the comparability of findings. With these combined efforts, CentiMarkers have the potential to revolutionize our understanding of AD and other diseases, expediting the development of effective treatments.

### Supporting information

Supplemental Figures

### Data Availability

All data produced are available online at:
https://dian.wustl.edu/our-research/clinical-trial/
https://adni.loni.usc.edu/

https://dian.wustl.edu/our-research/clinical-trial/edu/

https://adni.loni.usc.edu/

## Acknowledgements

We gratefully acknowledge the outstanding commitment of the participants, family members, and caregivers whose participation was critical to the success of the DIAN-TU trial. We thank the DIAN-TU study team for their exceptional dedication and accomplishments, which ensured the success of the trial. We thank the DIAN-EXR and DIAN-OBS study teams for their support on recruitment and commitment to family members. We appreciate the robust intellectual collaboration between the DIAN-TU investigators, participants and family members, F. Hoffmann-La Roche, Ltd./Genentech, and Eli Lilly and Company, the DIAN-TU Pharma Consortium (https://dian.wustl.edu/our-research/the-pharma-consortium/), the NIH, and regulatory representatives who were critical in making this study possible. We thank the Alzheimer’s Association, GHR Foundation, an anonymous organization, other industry partners (Avid Radiopharmaceuticals, a wholly-owned subsidiary of Eli Lilly and Company, Signant, and Cogstate), and regulatory representatives for their support. We also acknowledge Dr. Laurie Ryan from the National Institute on Aging for her key contributions in leadership and scientific guidance on this project.

## Consent Statement

The DIAN-TU study was conducted in accordance with the Declaration of Helsinki and the International Council for Harmonization and Good Clinical Practice guidelines and had ethics committee approval at each participating site. All participants provided written informed consent.

## Collaborators

### DIAN-TU Study Team

The DIAN-TU acknowledges the many individuals who have contributed to the DIAN-TU Trials including funding partners, leadership team, core leaders, project arm leaders, study sites and institutional study partners listed in the following pages.

### DIAN-TU Leadership Team

Randall Bateman, MD, Director and Principal Investigator Eric McDade, DO, Associate Director

David Clifford, MD, Associate Director and Medical Director Stephen Salloway, MD, Project Arm Leader, Gantenerumab Martin Farlow, MD, Project Arm Leader, Solanezumab

Lon Schneider, MD, Project Arm Leader

### DIAN-TU Core Leaders

Randall Bateman, MD, Administrative Core Leader, Washington University School of Medicine Anne Fagan, PhD, Biomarker Core Leader, Washington University School of Medicine Chengjie Xiong, PhD, Biostatistics Core Leader, Washington University School of Medicine Guoqiao Wang, PhD, Biostatistics Co-Core Leader, Washington University School of Medicine Jason Hassenstab, PhD, Cognition Core Leader, Washington University School of Medicine Alison Goate, DPhil, Genetics Core Leader, Mt. Sinai School of Medicine

Carlos Cruchaga, PhD, Genetics Core Leader, Washington University School of Medicine Tammie Benzinger, MD, Imaging Core Leader, Washington University School of Medicine Rick Perrin, MD, Neuropathology Core Leader, Washington University School of Medicine Jorge Llibre-Guerra, MD, MSc, Post-Doctoral Associate

Cliff Jack, MD, MRI, Mayo Clinic

Robert Koeppe, MD, PET Imaging, University of Michigan

Nigel Cairns, MD (retired), Neuropathology Core Leader, Washington University School of Medicine Peter Snyder, PhD, (former Cognition Core Leader), Brown University

## DIAN Expanded Registry (DIAN-EXR)

Eric McDade - Director

Randall Bateman – Associate Director

Jorge Llibre-Guerra – Post-Doctoral Associate

Ellen Ziegemeier - Senior Clinical Research Coordinator Jennifer Petranek - Clinical Coordinator I

Sarah Adams - Clinical Research Coordinator II

Susan Brandon - Clinical Research Coordinator IIDIAN-TU Staff

## DIAN-TU Faculty and Staff

Amanda Fulbright, Grant Specialist, Administration Core

Ron Hawley, IT Audiovisual and Interactive web designer, Administration Core Jacki Mallmann, Senior Grant Specialist, Administration Core

Karen McCann, Financial Accounting Assistant, Administration Core Julie Murphy, Accounting/Purchasing Assistant, Administration Core Anna Santacruz, Administrative Director, Administration Core Jeanette Schillizzi, Research Administrator, Administration Core Wendy Simpson, Financial Accounting Assistant, Administration Core Shannon Sweeney, Finance Analyst, Administration Core

Kelley Coalier, Lead Scientist, Biomarker Core Fatima Amtashar, QC Technician, Biomarker Core Sushila Sathyan, Archivist, Biomarker Core

Jennifer Stauber, Research Specialist, Biomarker Core Susan Mills, Director of Clinical Operations

Nicole Kelley, Associate Director of Clinical Operations

Stephanie Belyew, Clinical Trial Manager, Site, Vendor and GCP Management, Clinical Operations Angela Fuqua, Clinical Trial Manager, Clinical Scale, Drug Supply and Vendor Management, Clinical Operations

Inbal Meshulam, Clinical Trials Manager, Clinical Operations

Annette Stiebel, Clinical Trial Manager, Regulatory, Clinical Operations Jeanine Portell, Clinical Trial Manager, MRI Sites, Clinical Operations Bettina Bell, Clinical Trial Manager, Brain Donation, Clinical Operations Caryll Bentley, Contract Project Manager, Clinical Operations

Sharon Cirello, Senior Contract Data Manager, Clinical Operations

Nithyanjali Devarapalli, Contract Specialist Clinical Data Manager, Clinical Operations Arthur Gipson, Contract Specialist Clinical Data Manager, Clinical Operations

JaNeen Wisner, Contract Project Coordinator, Clinical Operations Tayona Mayhew, Data Management Specialist, Clinical Operations Zenobia Bridgewater, Clinical Research Coordinator, Clinical Operations Dana Burgdorf, Research Nurse Coordinator II, Clinical Operations

Molly Fitzgerald, Clinical Research Study Assistant I, Clinical Operations Erica Fowler, Clinical Research Coordinator I, Clinical Operations

Dottie Heller, Research Nurse Coordinator II, Clinical Operations Miranda Jany, Clinical Research Coordinator I, Clinical Operations Latoya Jones, Clinical Research Coordinator II, Clinical Operations Michelle Jorke, Clinical Research Coordinator II, Clinical Operations Paulette MacDougall, Research Nurse Coordinator II, Clinical Operations Eugene Rubin, QC, Clinical Operations

Jessi Smith, Senior Clinical Research Coordinator, Clinical Operations Mary Wolfsberger, Clinical Research Coordinator, Clinical Operations Andy Aschenbrenner, Assistant Professor, Cognition Core

Jennifer Smith, Professional Rater III, Cognition Core

Marisol Tahan, Clinical Research Coordinator, Cognition Core Theresa Butler, Research Lab Manager, Imaging Core

Lisa Cash, Senior Clinical Research Coordinator, Imaging Core Jon Christensen, Staff Scientist, Imaging Core

Aylin Dince, Research Assistant, Imaging Core

Tony Durbin, Senior Clinical Research Coordinator, Imaging Core Shaney Flores, Research Assistant, Imaging Core

Karl Friedrichsen, Research Assistant, Imaging Core Brian Gordon, Co-Investigator, Imaging Core

Russ Hornbeck, Project Manager, Imaging Core Nelly Joseph-Mathurin, Post-doc, Imaging Core Sarah Keefe, Research Assistant, Imaging Core Lakisha Lloyd, Research Coordinator, Imaging Core Laura Marple, Research Technician II, Imaging Core

Austin McCullough, Graduate Research Assistant, Imaging Core Stephanie Schultz, Pre-Doctoral Trainee, Imaging Core

Sally Schwarz, Co-Investigator, Imaging Core Yi Su, Co-Investigator, Imaging Core

Andrei Vlassenko, Co-Investigator, Imaging Core Qing Wang, Post-doc, Imaging Core

Jinbin Xu, Co-Investigator, Imaging Core

Erin Franklin, Research Coordinator, Neuropathology Core

## Eli Lilly and Company and Avid Radiopharmaceuticals, a wholly owned subsidiary of Eli Lilly and Company

Eli Lilly and Company Avid Radiopharmaceuticals John Sims, MD, Eli Lilly and Company. Michael Devous

Karen Holdridge, MPH, Eli Lilly and Company. Erica Elephant Cheryl Brown, BS, Eli Lilly and Company. Laura Harper Roy Yaari, MD, Eli Lilly and Company Marybeth Howlett Isabella Velona, Clinical Trial Project Manager Mark Mintun Scott Andersen, Biostatistician Michael Pontecorvo

Michele Mancini, GPS

Brian Willis, PK/PD Project Leader

Jillian Venci Fuhs, Global Regulatory Affairs Julie Bush, Product Delivery

Shamrock Garrett, Sr. CTMA

Traci Peddie, Data Sciences & Solutions Natalie Vantwoud, Data Management Barbara Lightfoot-Owens, Medical Writer John Brad-Holmes, Central Lab

## Former Team Members

Phyllis Ferrell Barkman, Russ Barton, Lauren Brunke, Robert Dean, Deanilee Deckard, Ann Catherine Downing, Ganapathy Goppalrathnam, David Henley, Janice Hitchcock, Sonia Nijampatnam,Tracie Peddie, Melissa Pugh, Tami Jo Rayle, Shiloh Scott, James Senetar, Gopalan Sethuraman, Eric Siemers, Brian Steuerwald, Connie Tong, Jim Vandergriff

## F. Hoffman-LaRoche Team

Monika Baudler, LifeCycle Leader

Rachelle Doody, Global Head of Neurodegneration Paul Delmar, Principal Statistical Scientist

Carsten Hofmann, Clinical Pharmacologist Michaela Jahn, Global Biometrics Team Leader Geoff Kerchner, Global Development Leader

Gregory Klein, Biomarker Experimental Medicine Leader Smiljana Ristic, Associate Group Medical Director Alison Searle, Operations Program Leader

Marco Sonderegger, Technical Development Leader Roz Sutton, EU Regulatory Partner

Janette Turner, US Regulatory Partner Jaku Wojtowicz, Safety Science Director Susan Yule, Global Regulatory Leader

## Former Team Members

Elizabeth Ashford, Operations Program Leader; Bogdon Balas, Safety Science Leader; Estelle Vester- Blokland, LifeCycle Leader; Stephanie Capo-Chichi, EU Regulatory Partner; David Agnew, Global Study Manager; Ernest Dorflinger, Translational Medicine Leader; Efe Egharevba, Global Study Manager; Christelle Laroche, Technical Development Leader; Isabelle Bauer Dauphin, Technical Development Leader; Rob Lasser, Global Development Leader; Ferenc Martenyi, Global Development Leader; Glenn Morrison, Global Development Leader; Tania Nikolcheva, Biomarker Experimental

Medicine Leader; Michael Rabbia, Statistical Scientist; Juha Savola, Project Leader; Janice Smith, Clinical Science Leader; Dietmar Volz, Statistical Scientist

## DIAN-TU DSMB Members

Gary Cutter, PhD, DSMB Chairperson, University of Alabama at Birmingham Steve Greenberg, MD, PhD, Massachusetts General Hospital, Boston

Scott Kim, MD, PhD, National Institutes of Health, Bethesda David Knopman, MD, Mayo Clinic, Rochester

Willis Maddrey, MD (retired), UT Southwestern, Dallas Kristine Yaffe, MD, University of California, San Francisco

Karl Kieburtz, MD, PhD, (DSMB Chairperson, retired) University of Rochester, NY Allan Levey, MD, PhD (retired), Emory University, Atlanta

## DIAN-TU Therapy Evaluation Committee

Randall Bateman, MD, Chair, Washington University School of Medicine, St. Louis Eric McDade, DO, Co-Chair, Washington University School of Medicine, St. Louis Paul Aisen, MD, Alzheimer’s Therapeutic Research Institute, USC

Jasmeer Chhatwal, MD, PhD, MMSc, Massachusetts General Hospital, Harvard Medical School David Clifford, MD, Washington University School of Medicine, St. Louis

David Cribbs, MD, UC Irvine, CA

Nick Fox, MD, FRCP, FMedSci, Dementia Research Centre

Serge Gauthier, Serge Gauthier, CM, MD, FRCPC, Director, AD & Related Disorders Unit McGill Centre for Studies in Aging

David Holtzman, MD, Washington University School of Medicine

Matthias Jucker, PhD, Hertie Institute for Clinical Brain Research, DZNE, Germany Jeff Kelly, MD, Scripps University, California

Virginia Lee, PhD, University of Pennsylvania, Perelman School of Medicine Simon Mead, FRCP, PhD, Institute of Prion Diseases, London

Cath Mummery, PhD, FRCP, Dementia Research Centre, London Erik Musiek, MD, PhD, Washington University School of Medicine Erik Roberson, MD, PhD, University of Alabama

Mathias Staufenbiel, PhD, Hertie Institute for Clinical Brain Research, DZNE, Tubingen, Germany Robert Vassar, PhD, Northwestern University, IL

## Former TEC Members

Bart DeStrooper, PhD; William Klunk, MD, PhD; Cynthia Lemere, MD; John C. Morris, MD

## DIAN Clinical Trials Committee (CTC) Members

Randall Bateman, Washington University in St. Louis School of Medicine John Morris, Washington University in St. Louis School of Medicine Chengjie Xiong, Washington University in St. Louis School of Medicine Denise Heinrichs, DIAN Family Representative

John Ringman University of California, Los Angeles

Laurie Ryan, Division of Science, National Institute on Aging Neil Buckholtz, National Institute on Aging

Reisa Sperling, Director, Center for Alzheimer Research and Treatment Stephen Salloway, Butler Hospital

Paul Aisen, University of California, San Diego

Anna Santacruz, Washington University in St. Louis School of Medicine Gabrielle Strobel, Alzheimer Research Forum

Bill Klunk, University of Pittsburgh

William Thies, Alzheimer’s Association

Anne Fagan, Washington University in St. Louis School of Medicine Mark Mintun, Washington University in St. Louis School of Medicine Natalie Ryan, University College London

Virginia Buckles, Washington University in St. Louis School of Medicine David Hawver, Food and Drug Administration

Martin Farlow, Indiana University

Maritza Ciliberto, DIAN Family Representative Ralph Martins, Edith Cowan University Jennifer Williamson, Columbia University

## Study Sites

**Australia**: Neuroscience Research Australia - W Brooks, MJ Fulham, J Bechara, D Foxe; Australian Alzheimer’s Research Foundation – R Clarnette, N Reynders, P Mather; University of Melbourne – C Masters, C Rowe, B Clinch, D Baxendale

**Canada**: McGill University – S Gauthier, P Rosa-Neto, C Mayhew, L Robb; University of British Columbia – R Hsiung, D Worsley, M Assaly, E Nicklin; Sunnybrook Research Institute – M Masellis, K Sharp, S Hetherington

**France:** Hopital Charles Nicolle – D Wallon, D Hannequin, A Morin, A Zarea, E Gerardin, P Bohn, M Chastan, P Vera, M Colnot, N Donnadieu, M Quillard-Muraine, C Bergot, S Jourdain; Groupe Hospitalier Pitie-Salpetriere – B Dubois, M Habert, N Younsi; Hopital Pierre Wertheimer – M Formaglio, D Lebars, N El Kfif, A Jullien; Hopital Purpan – J Pariente, P Payoux, C Thalamas, A Driff, E Pomies, P Gauteul; Hôpital Roger Salengro – F Pasquier, A Rollin-Sillaire, F Semah, L Breuilh, M Laforce; Orsay Imaging – M Bottlaender

**Spain:** Hospital Clinic i Provincial de Barcelona – R Sanchez-Valle, M Balasa, A Lladó, B Bosch, N Bargalló, I Banzo, A Perisinotti

**United Kingdom**: University College London Hospital – C Mummery, I Kayani, J Douglas, M Grilo **United States:** Washington University School of Medicine – BJ Snider, T Benzinger, W Sigurdson, T Donahue, P Kelly; Emory University – J Lah, C Meltzer, G Schwartz, P Vaughn, L Piendel; University of Pittsburgh – S Berman, J Mountz, L Macedonia, S Ikonomovic, S Goldberg, E Weamer, J Ruskiewiecz, S Hegedus, L Tarr, T Potter, G Valetti; University of Alabama at Birmingham – E Roberson, D Geldmacher, M Love, A Watkins, L Ashley; Indiana University – J Brosch, A Kohn, N McClaskey, J Buck, J Fletcher; Butler Hospital – G Surti, R Noto, C Bodge, W Menard; University of Puerto Rico – I Jimenez Valazquez, J Diaz, K Aleman; Yale University – C van Dyck, M Chen, N Diepenbrock, A Mecca, S Good; University of California San Diego – D Galasko, C Hoh, D Szpak, S Peackock; University of Washington – S Jayadev, D Lewis, Y Tutterow

## DIAN-TU Collaborators and Advisers

John Morris, MD, Senior Advisor, Washington University School of Medicine, St. Louis, MO David Holtzman, MD, Senior Advisor, Washington University School of Medicine, St. Louis, MO Laura Swisher, MS, Deputy Director, Washington University School of Medicine, St. Louis, MO Alisha Daniels, MD, MHA, Executive Director, DIAN, Washington University School of Medicine, St. Louis, MO

Janice Hitchcock, PhD, Hitchcock Regulatory Consulting Inc. Thomas Bird, MD, University of Washington, Seattle

Dennis Dickinson, MD, Mayo Clinic, Jacksonville, FL

M. Marsel Mesulam, MD, Cognitive Neurology and Alzheimer’s Disease Center, Northwestern University, Chicago, IL

Cornelia Kamp, MBA, University of Rochester, New York Ron Thomas, PhD, ADCS, University of California, San Diego

Paul Aisen, MD, ADCS, University of Southern California, Los Angeles

## DIAN Observational Study Site Investigators

James Noble, MD, Columbia University, New York Martin Farlow, MD, Indiana University, Indianapolis, IN

Jasmeer Chhatwal, MD, PhD, Brigham and Women’s Hospital-Massachusetts GH, Charlestown, MA Stephen Salloway, MD, Butler Hospital, Warren Alpert School of Medicine, Brown University

Sarah Berman, MD, PhD, University of Pittsburgh, PA Gregg Day, MD, Mayo Clinic Jacksonville, FL

Hiroyuki Shimada, MD, PhD, Osaka City University, Japan Takeshi Ikeuchi, MD, PhD, Brain Research Institute, Nigata, Japan Kazushi Suzuki, MD, PhD, The University of Tokyo, Japan

Peter Schofield, PhD, DSc, Neuroscience Research Australia, Sydney

Ralph Martins, BSc, PhD, Edith Cowan University, Nedlands, Western Australia

Nick Fox, MD, FRCP, FMedSci, Dementia Research Centre, University College London, United Kingdom

Johannes Levin, MD, PhD, German Center for Neurodegenerative Diseases (DZNE), Munich, Germany Mathias Jucker, PhD, German Center for Neurodegenerative Diseases (DZNE), Tubingen, Germany Raquel Sanchez Valle, MD, Hospital Clinic i Provincial de Barcelona, Spain

Patricio Chrem, MD, Fundación para la Lucha contra las Enfermedades Neurológicas de la Infancia (FLENI), Buenos Aires, Argentina

## DIAN EXR Referring Clinicians, Researchers and Partner Sites

Neelum T. Aggarwal, MD, Rush University Medical Center, Chicago IL

Tom Ala, Center for Alzheimer’s Disease and Related Disorders, Southern Illinois University School of Medicine

Thomas Bird, University of Washington, Seattle

Sandra E. Black, Sunnybrook Health Sciences Centre, University of Toronto, Canada William J. Burke, MD, Banner Alzheimer’s Institute

Cynthia M. Carlsson, MD, MS, University of Wisconsin School of Medicine and Public Health Andrew Frank M.D. B.Sc.H. F.R.C.P.(C), Bruyere Continuing Care, Ottawa, Ontario, Canada James E. Galvin, MD, MPH, Charles E. Schmidt College of Medicine, Florida Atlantic University Alvin C Holm, MD, Bethesda Hospital, St. Paul, MN

John S.K. Kauwe, Brigham Young University David Knopman MD, Mayo Clinic, Rochester MN

Sarah Kremen, MD, University of California, Los Angeles Alan J. Lerner, University Hospitals Cleveland Medical Center Barry S. Oken, MD, PhD, Oregon Health & Science University Hamid R. Okhravi, Eastern Virginia Medical School

Ronald C. Petersen, Mayo Clinic, Rochester, MN Aimee L. Pierce, MD, University of California Irvine

Marsha J. Polk, MED, University of Texas Health Science Center at San Antonio John M. Ringman, MD, MS, University Southern California

Peter St. George Hyslop, MD, FRS, FRSC, FRCPC, University of Toronto Sanjeev N. Vaishnavi, MD, PhD, University of Pennsylvania

Sandra Weintraub, Northwestern University Feinberg School of Medicine, IL

