## Supplemental Figures for "The CentiMarker Project: Standardizing Quantitative Alzheimer’s disease Fluid Biomarkers for Biologic Interpretation"

### Supplemental Materials

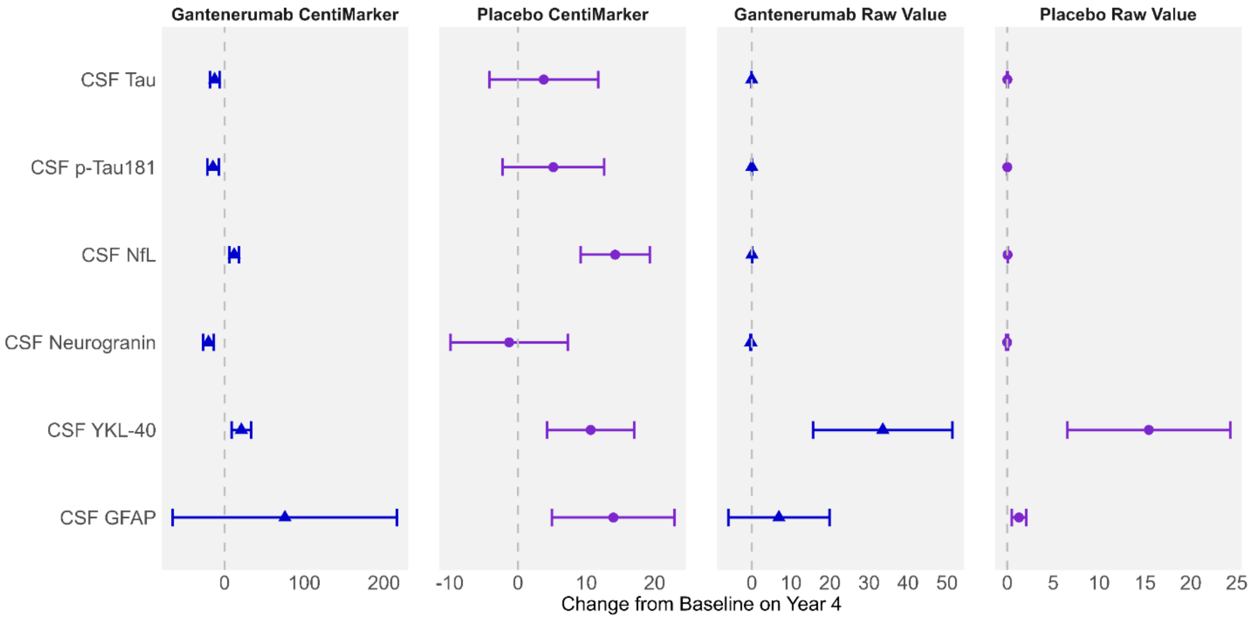


**Supplement Figure 1**: Utilizing CentiMarkers facilitates the interpretation and comparison across biomarkers by converting them to a similar scale, while using raw values is more difficult to compare as units are different and not scaled to disease ranges. Estimated mean change from baseline in CentiMarkers with 95% confidence intervals for the treatment and shared placebo groups using MMRM analyses in the DIAN-TU-001 trial of gantenerumab. These results demonstrate the magnitude of disease normalization compared to normal (CM 0) vs. fully abnormal (CM 100) states. Raw values are in the unit of ng/mL

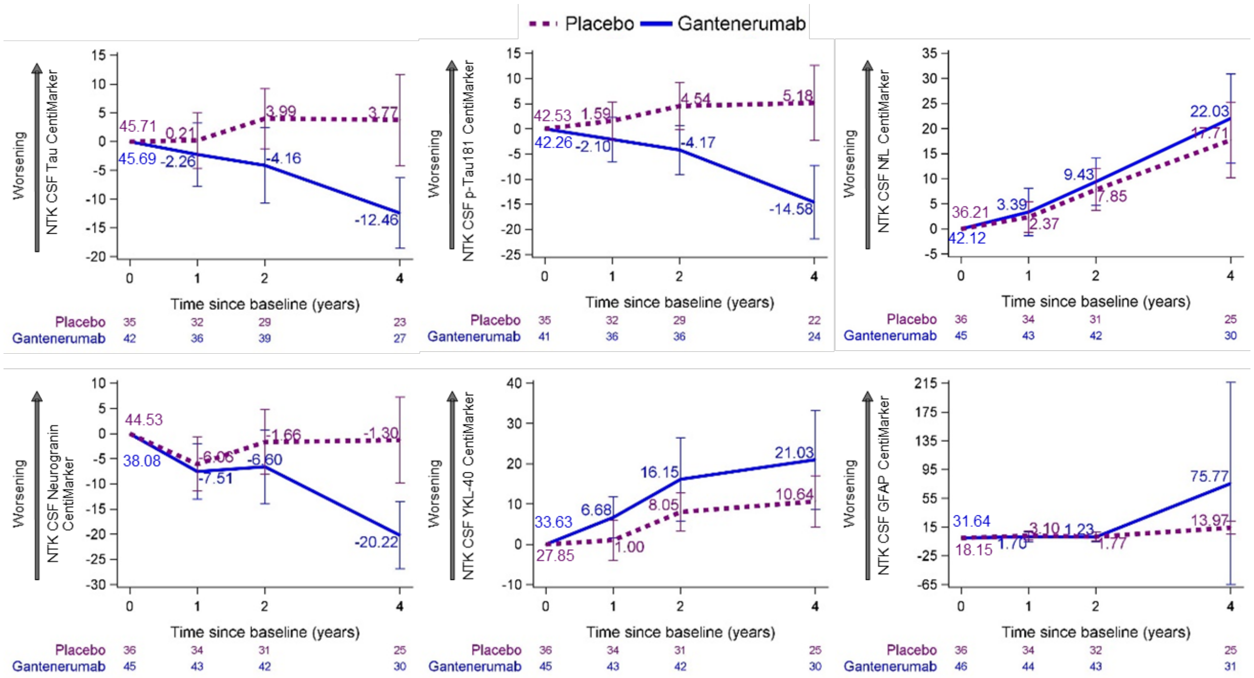


**Supplemental Figure 2**: Estimated mean change from baseline in CentiMarkers with 95% confidence intervals for the gantenerumab and the placebo groups using MMRM analyses in the DIAN-TU-001 trial. These results demonstrate the magnitude of disease normalization compared to normal (CM 0) vs. fully abnormal (CM 100) states.
